# Assessing glucose-6-phosphate dehydrogenase (G6PD) during COVID-19 requires caution: evidence on the impact of the infection upon enzyme activity

**DOI:** 10.1101/2022.08.25.22279199

**Authors:** Maria Gabriela de Almeida Rodrigues, Gisely Cardoso de Melo, Ádila Liliane Barros Dias, Marco Aurélio Sartim, Mariana Simão Xavier, Rebeca Linhares Abreu Netto, Fernando Fonseca Almeida Val, Djane Clarys Baía-da-Silva, José Diego Brito-Sousa, Wuelton Marcelo Monteiro, Marcus Vinicius Guimarães de Lacerda, Vanderson de Souza Sampaio

## Abstract

Glucose-6 phosphate dehydrogenase deficiency (G6PDd) was suggested as a risk factor of severity in patients with COVID-19. In this article, we assessed the influence of G6PDd on the infection, severity, and clinical progression of patients with COVID-19. This prospective cohort study included adult participants (≥18 years old) who had clinical and/or radiological COVID-19 findings or positive RT-PCR results. Epidemiological and clinical data were extracted from electronic medical records. G6PD activity was measured in SD Biosensor STANDARD G6PD® equipment at admission and one year after discharge. Samples were genotyped for the three most common single nucleotide polymorphisms (SNPs) for G6PDd in the Brazilian Amazon s1050828, rs1050829 and rs5030868, corresponding to G6PD African A-(G202A, A376G), G6PD African A+(A376G) and G6PD Mediterranean(C563T), respectively. Seven hundred fifty-three patients were included, of which 123 (16.3%) were G6PDd. The G6PDd group had a higher mean hemoglobin, and lower values of C-reactive protein and leukocytes at admission. There was no association between G6PDd and COVID-19 severity, considering that the frequency of G6PDd who needed to be hospitalized (1.9%) or demanding invasive mechanical ventilation (16.0%) or died (21.1%) was lower than G6PD normal patients. Only 29 out of 116 (25%) participants carried the African genotype. Out of 30 participants tested as G6PDd during disease, only 11 (36.7%) results agreed one year after discharge. In conclusion, caution must be taken when G6PDd screening in patients with acute COVID-19.

## INTRODUCTION

COVID-19, an infectious disease caused by the SARS-CoV-2 infection, is the most critical public health problem in the world, and its impact is evident due to its rapid spread [1]. SARS-CoV-2 infection can be asymptomatic in most cases, but ca be progress to severe illness, characterized by pneumonia, severe respiratory syndrome (SARS), and multiorgan failure, with high associated mortality rates [2–4]. Older people, people with chronic diseases, including diabetes, cardiovascular diseases, cancer, and chronic respiratory diseases, are more likely to evolve to severity and death [5–10]. Biochemical factors that potentially predispose individuals to the development of severe and excessive complications associated with the disease are unclear [11]. Some reports of glucose-6-phosphate dehydrogenase enzyme deficiency (G6PDd), an important enzyme involved in the hexose monophosphate pathway, associated with COVID-19 are found in the literature [12–18].

G6PDd is a genetic condition that affects approximately 400 million people worldwide [19,20], being an inherited enzyme deficiency linked to the X chromosome caused by mutations in this gene encoding the enzyme G6PD [21]. In erythrocytes, G6PD is the only source of enzyme activity that protects the cell against oxidative stress [22]. G6PDd is related to neonatal hyperbilirubinemia, chronic hemolytic anemia, acute hemolysis, and risk of acute hemolytic anemia [23–25]. Although most affected individuals are asymptomatic during lifetime, exposure to oxidative stressors such as certain drugs and other substances, or infections can elicit acute hemolytic anemia [26,27]. Studies suggest that the pathogenesis of SARS-COV-2 involves the overproduction of Reactive Oxygen Species (ROS) [28,29]. The production of excessive ROS is critical in individuals with G6PDd because the antioxidant system is compromised [30], and this can contribute to the progression and severity of respiratory disease [29,31–33]. Thus, G6PDd was suggested as a health condition that may increase the risk of severity and mortality in people with COVID-19 [34,35].

Observational studies report that G6PDd patients are more hospitalized than normal G6PD patients (G6PDn) [14,36], in addition, among those hospitalized requiring supplemental oxygen, the G6PDd had worse hematological indices and the G6PDd group had a prolonged PaO2/FiO2 ratio and longer days on mechanical ventilation, indicating the severity of the pneumonia [37]. Vick [38]suggests that COVID-19 infection may be a trigger for hemolysis and coagulopathy in patients with G6PDd, explaining the stroke symptoms common to severe COVID-19 conditions[39,40]. Kumar et al. [41], however, showed that the occurrence of non-invasive ventilation, intubation or death – all of which are indicative of severe COVID-19, are not significantly different in hospitalized patients with COVID-19 with and without G6PDd. Here we assessed the influence of G6PDd on clinical progression of patients with COVID-19 and compared G6PD activity during and post COVID-19. The results presented here show the potential modulation of enzymatic activity on COVID-19.

## MATERIALS AND METHODS

### Study design and participants

This cohort study included adult (≥18 years old) patients seen at the *Hospital e Pronto-Socorro Delphina Rinaldi Abdel Aziz* and *Unidade de Pronto Atendimento (UPA) Campos Sales* located in Manaus, Amazonas, Brazil, between March 23 to 24 May 2020. Manaus is the most populous municipality in Amazonas, with an estimated population of 2.2 million inhabitants, and has become an important transmission site for SARS-CoV-2 in Brazil [42]. The participants were included if they had clinical and/or radiological suspicion of COVID-19 (history of fever AND any respiratory symptom, e.g., cough or dyspnea AND/OR ground-glass opacity OR pulmonary consolidation on CT scan) or RT-PCR positive.

### Data collection

Epidemiological, demographic, clinical, laboratory, treatment, and outcome data were extracted from electronic medical records using a standardized data collection and then transferred to an electronic database (REDCap). Clinical parameters were measured daily by the hospital’s clinical staff from day one until discharge or death, and daily patient information from the first visit to 28 days or until death was collected. One year later, patients who survived were contacted and who agreed to have a new sample collected were evaluated for G6PD enzymatic activity.

### Laboratory procedures

Laboratory tests were carried out according to the hospital’s routine and at the doctor’s discretion. Hematology and biochemistry analyses were performed in automated machines. COVID-19 diagnosis was performed in nasopharyngeal and oropharyngeal swabs sample using molecular techniques, according to the protocol developed by the US Centers for Disease Control and Prevention (CDC/USA), updated on March 15, 2020 (30). Swab specimens were collected on days 1, 5 and 7. In D1 and one year from infection, the measurement of the G6PD enzyme activity and G6PD and hemoglobin occurred using the SD Biosensor STANDARD G6PD® equipment (31). According to the activity of the G6PD enzyme, patients were divided into two groups: deficient G6PD (G6PDd) [activity<6] and normal G6PD (G6PDn) [activity≥6] [43].

Samples were genotyped for the three most common single nucleotide polymorphisms (SNPs) for G6PDd in the Brazilian Amazon [44,45] s1050828, rs1050829 and rs5030868, corresponding to G6PD African A-(G202A, A376G), G6PD African A+(A376G) and G6PD Mediterranean(C563T), respectively [46]

### Definitions

The minimum values of hemoglobin and lymphocytes were considered laboratory parameters of severity, as well as the maximum values of C-reactive protein, total, direct, indirect bilirubin, AST, ALT, LDH, and D-dimer at the follow-up. The clinical progression of COVID-19 was assessed considering hospitalization in the ward and ICU, need for invasive and non-invasive mechanical ventilation, length of stay in the hospital and ICU, time until admission to the ICU, and death. According to hospital protocol, invasive mechanical ventilation (IVM) was recommended when PaO2 / FiO2 <150.

### Statistical analysis

The proportions between groups were compared using the chi-square test for categorical variables. The mean and standard deviation, as well as the median and interquartile range were calculated for continuous variables and the comparison between groups was performed using ANOVA and Wilcoxon rank-sum, accordingly. To compare the laboratory parameters between the groups, the Wilcoxon rank-sum test was performed. Comparison of G6PD activity during and after COVID-19 was performed by Wilcoxon matched pair signed classification test. All analyzes were performed using the Stata statistical package (v.17).

### Ethical considerations

We used data from medical records of patients included in clinical trials conducted by the research team. These trials were approved by the Brazilian Committee of Ethics in Human Research for their respective original objectives (CAAE: 30152620.1.0000.0005, 30504220.5.0000.0005, and 30615920.2.0000.0005). All participants signed an informed consent form at the time of inclusion in the primary studies or, when unable, a family member performed the consent process. Participant’s sensitive data were not available to avoid identification. All measures were taken to ensure the participants’ confidentiality in accordance with the good clinical practice.

## RESULTS

A total of 753 participants were included, of which 123 (16.3%) were G6PDd in G6PD quantitative test. Demographic and clinical characteristics at admission are shown in Table 1. Hemoglobin concentration was significantly higher for the G6PDd group (13.2, P = 0.001). The G6PDn group had higher values of C-reactive protein (79.5, P = 0.009) and leukocytes (11,443.1, P <0.001) at admission. A higher proportion of G6PDn patients was at invasive mechanical ventilation (25.9%, P = 0.004) and hospitalized (70.4%, P = 0.002) at admission. About 45% of patients used chloroquine combined with antibiotics. The frequency of medication used previously or during hospitalization by group is shown in Supplementary Table 1 and 2, respectively. The use of insulin (31.2%, P = 0.027) and vasoactive amines (30.6%, P = 0.012) was significantly higher in the G6PDn group at follow-up. Regarding clinical outcomes, 29.8% of the study population died, with the proportion of death significantly higher in the G6PDn group (P = 0.022). Other outcomes showed similar proportions between groups (Table 2).

**Table 1.**
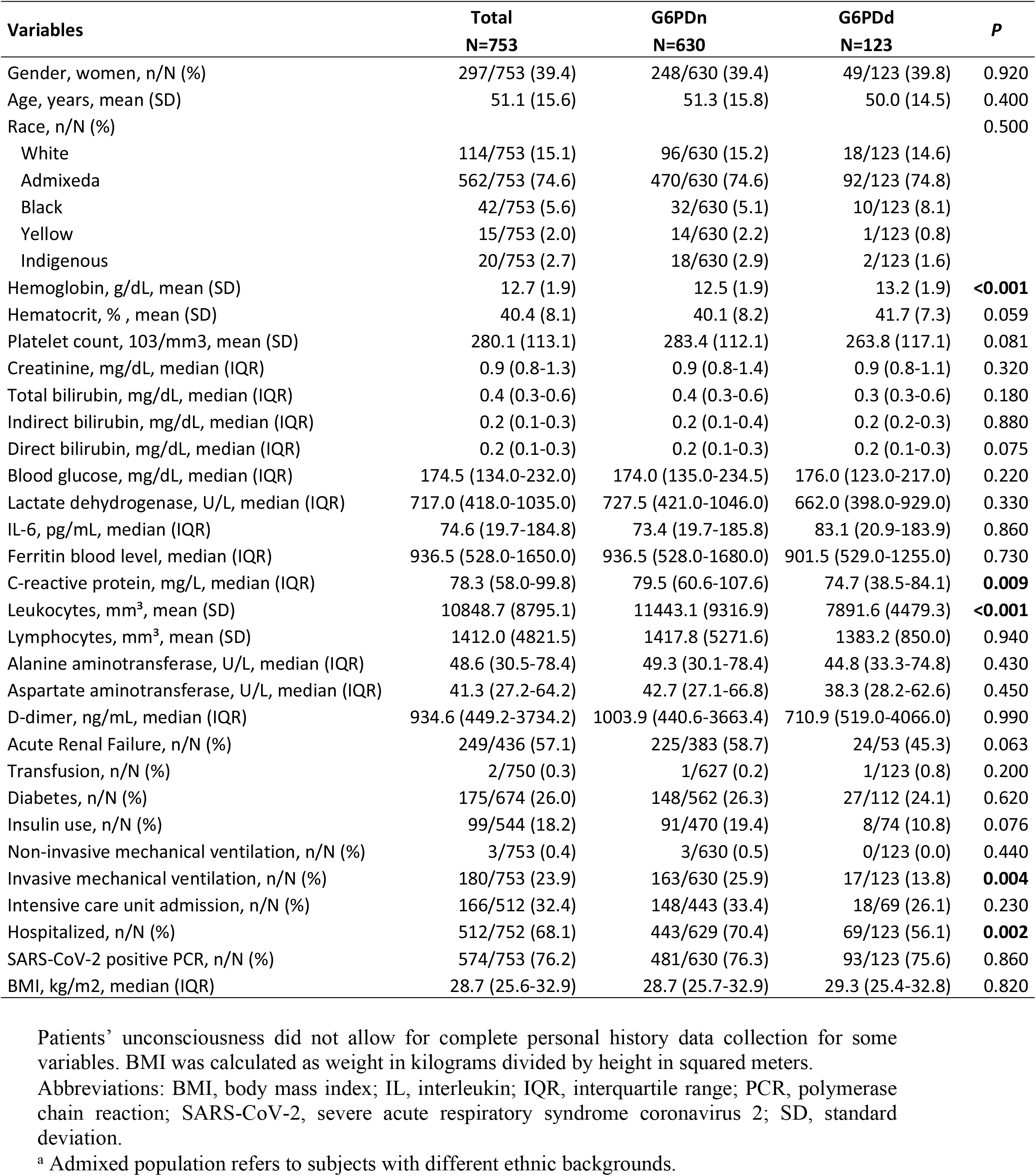
Demographic and clinical characteristics of the patients evaluated.

| Variables | Total<br>N=753 | G6PDn<br>N=630 | G6PDd<br>N=123 | P |
| --- | --- | --- | --- | --- |
| Gender, women, n/N (%) | 297/753 (39.4) | 248/630 (39.4) | 49/123 (39.8) | 0.920 |
| Age, years, mean (SD) | 51.1 (15.6) | 51.3 (15.8) | 50.0 (14.5) | 0.400 |
| Race, n/N (%) |  |  |  | 0.500 |
| White | 114/753 (15.1) | 96/630 (15.2) | 18/123 (14.6) |  |
| Admixed<sup>a</sup> | 562/753 (74.6) | 470/630 (74.6) | 92/123 (74.8) |  |
| Black | 42/753 (5.6) | 32/630 (5.1) | 10/123 (8.1) |  |
| Yellow | 15/753 (2.0) | 14/630 (2.2) | 1/123 (0.8) |  |
| Indigenous | 20/753 (2.7) | 18/630 (2.9) | 2/123 (1.6) |  |
| Hemoglobin, g/dL, mean (SD) | 12.7 (1.9) | 12.5 (1.9) | 13.2 (1.9) | **<0.001** |
| Hematocrit, %, mean (SD) | 40.4 (8.1) | 40.1 (8.2) | 41.7 (7.3) | 0.059 |
| Platelet count, 103/mm<sup>3</sup>, mean (SD) | 280.1 (113.1) | 283.4 (112.1) | 263.8 (117.1) | 0.081 |
| Creatinine, mg/dL, median (IQR) | 0.9 (0.8-1.3) | 0.9 (0.8-1.4) | 0.9 (0.8-1.1) | 0.320 |
| Total bilirubin, mg/dL, median (IQR) | 0.4 (0.3-0.6) | 0.4 (0.3-0.6) | 0.3 (0.3-0.6) | 0.180 |
| Indirect bilirubin, mg/dL, median (IQR) | 0.2 (0.1-0.3) | 0.2 (0.1-0.4) | 0.2 (0.2-0.3) | 0.880 |
| Direct bilirubin, mg/dL, median (IQR) | 0.2 (0.1-0.3) | 0.2 (0.1-0.3) | 0.2 (0.1-0.3) | 0.075 |
| Blood glucose, mg/dL, median (IQR) | 174.5 (134.0-232.0) | 174.0 (135.0-234.5) | 176.0 (123.0-217.0) | 0.220 |
| Lactate dehydrogenase, U/L, median (IQR) | 717.0 (418.0-1035.0) | 727.5 (421.0-1046.0) | 662.0 (398.0-929.0) | 0.330 |
| IL-6, pg/mL, median (IQR) | 74.6 (19.7-184.8) | 73.4 (19.7-185.8) | 83.1 (20.9-183.9) | 0.860 |
| Ferritin blood level, median (IQR) | 936.5 (528.0-1650.0) | 936.5 (528.0-1680.0) | 901.5 (529.0-1255.0) | 0.730 |
| C-reactive protein, mg/L, median (IQR) | 78.3 (58.0-99.8) | 79.5 (60.6-107.6) | 74.7 (38.5-84.1) | **0.009** |
| Leukocytes, mm<sup>3</sup>, mean (SD) | 10848.7 (8795.1) | 11443.1 (9316.9) | 7891.6 (4479.3) | **<0.001** |
| Lymphocytes, mm<sup>3</sup>, mean (SD) | 1412.0 (4821.5) | 1417.8 (5271.6) | 1383.2 (850.0) | 0.940 |
| Alanine aminotransferase, U/L, median (IQR) | 48.6 (30.5-78.4) | 49.3 (30.1-78.4) | 44.8 (33.3-74.8) | 0.430 |
| Aspartate aminotransferase, U/L, median (IQR) | 41.3 (27.2-64.2) | 42.7 (27.1-66.8) | 38.3 (28.2-62.6) | 0.450 |
| D-dimer, ng/mL, median (IQR) | 934.6 (449.2-3734.2) | 1003.9 (440.6-3663.4) | 710.9 (519.0-4066.0) | 0.990 |
| Acute Renal Failure, n/N (%) | 249/436 (57.1) | 225/383 (58.7) | 24/53 (45.3) | 0.063 |
| Transfusion, n/N (%) | 2/750 (0.3) | 1/627 (0.2) | 1/123 (0.8) | 0.200 |
| Diabetes, n/N (%) | 175/674 (26.0) | 148/562 (26.3) | 27/112 (24.1) | 0.620 |
| Insulin use, n/N (%) | 99/544 (18.2) | 91/470 (19.4) | 8/74 (10.8) | 0.076 |
| Non-invasive mechanical ventilation, n/N (%) | 3/753 (0.4) | 3/630 (0.5) | 0/123 (0.0) | 0.440 |
| Invasive mechanical ventilation, n/N (%) | 180/753 (23.9) | 163/630 (25.9) | 17/123 (13.8) | **0.004** |
| Intensive care unit admission, n/N (%) | 166/512 (32.4) | 148/443 (33.4) | 18/69 (26.1) | 0.230 |
| Hospitalized, n/N (%) | 512/752 (68.1) | 443/629 (70.4) | 69/123 (56.1) | **0.002** |
| SARS-CoV-2 positive PCR, n/N (%) | 574/753 (76.2) | 481/630 (76.3) | 93/123 (75.6) | 0.860 |
| BMI, kg/m<sup>2</sup>, median (IQR) | 28.7 (25.6-32.9) | 28.7 (25.7-32.9) | 29.3 (25.4-32.8) | 0.820 |

Patients' unconsciousness did not allow for complete personal history data collection for some variables. BMI was calculated as weight in kilograms divided by height in squared meters.
Abbreviations: BMI, body mass index; IL, interleukin; IQR, interquartile range; PCR, polymerase chain reaction; SARS-CoV-2, severe acute respiratory syndrome coronavirus 2; SD, standard deviation.
<sup>a</sup> Admixed population refers to subjects with different ethnic backgrounds.

**Table 2.**
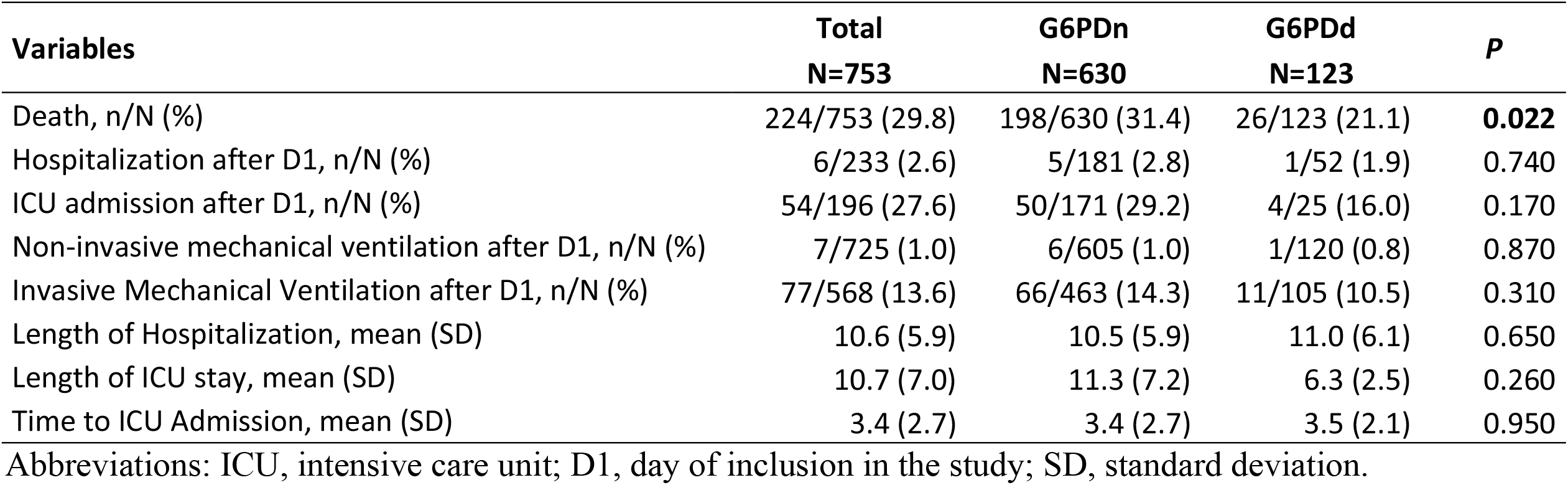
Clinical characteristics throughout the follow-up.

| **Variables** | **Total**<br>**N=753** | **G6PDn**<br>**N=630** | **G6PDd**<br>**N=123** | **P** |
| --- | --- | --- | --- | --- |
| Death, n/N (%) | 224/753 (29.8) | 198/630 (31.4) | 26/123 (21.1) | **0.022** |
| Hospitalization after D1, n/N (%) | 6/233 (2.6) | 5/181 (2.8) | 1/52 (1.9) | 0.740 |
| ICU admission after D1, n/N (%) | 54/196 (27.6) | 50/171 (29.2) | 4/25 (16.0) | 0.170 |
| Non-invasive mechanical ventilation after D1, n/N (%) | 7/725 (1.0) | 6/605 (1.0) | 1/120 (0.8) | 0.870 |
| Invasive Mechanical Ventilation after D1, n/N (%) | 77/568 (13.6) | 66/463 (14.3) | 11/105 (10.5) | 0.310 |
| Length of Hospitalization, mean (SD) | 10.6 (5.9) | 10.5 (5.9) | 11.0 (6.1) | 0.650 |
| Length of ICU stay, mean (SD) | 10.7 (7.0) | 11.3 (7.2) | 6.3 (2.5) | 0.260 |
| Time to ICU Admission, mean (SD) | 3.4 (2.7) | 3.4 (2.7) | 3.5 (2.1) | 0.950 |

Abbreviations: ICU, intensive care unit; D1, day of inclusion in the study; SD, standard deviation.

The laboratory markers throughout the follow-up are shown in Supplementary Figure 1. The G6PDd group presented higher values, especially in D14, even though standard deviations overlap, considering direct, indirect, and total bilirubin. These values are similar between groups up to D28. The minimum values of hemoglobin (Z = -3.931; P < 0.001) and lymphocytes (Z = -3.011; P = 0.003) were statistically different between groups (Figure 1 A, C). Maximum direct bilirubin was higher in the normal G6PD group (Z = 2.098; P = 0.036) (Figure 1 F). The other parameters evaluated showed no significance between groups (Figure 1 B, D, E, G-J).

**Figure 1.**
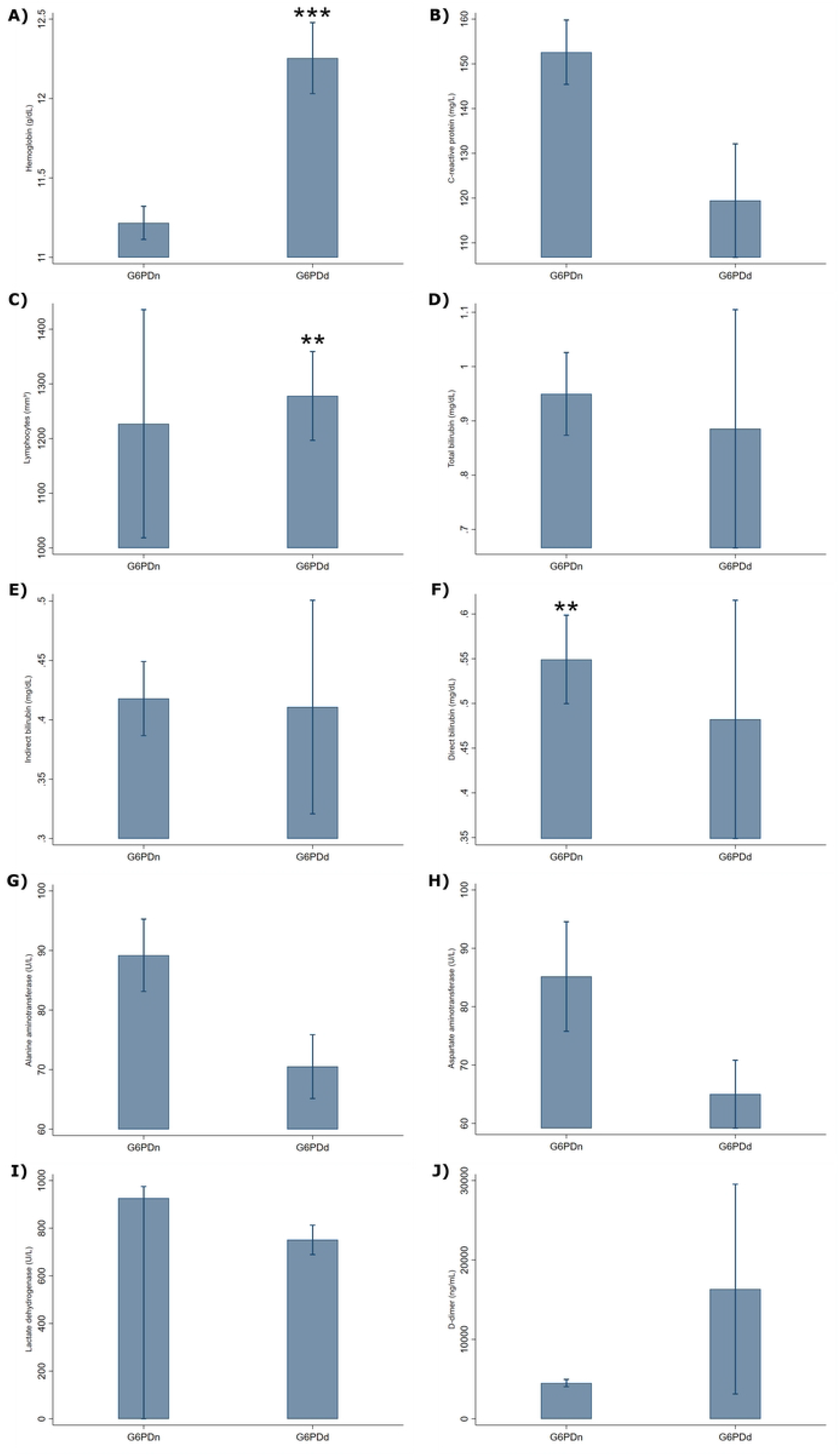
Mean of the severity laboratory parameters evaluated. Hemoglobin (A); C-reactive protein (B); Lymphocytes (C); Total bilirubin (D); Indirect bilirubin (E); Direct bilirubin (F); Alanine aminotransferase (G); Aspartate aminotransferase (H); Lactate dehydrogenase (I); D-dimer (J). Groups compared by Wilcoxon rank-sum test, with p<0.001(***) and p<0.05(**).

Out of the total G6PDd participants, 116 were genotyped, with 29 (25%) presenting a known deficiency variant. Fifteen out of 116 (12.9%) were classified as African+, 14 African-(12.1%), and 87 as wild genotypes (75%). One year from infection, 30 selected participants, previously classified as G6PDd by phenotype were retested, from which only 11 (36.7 %) were confirmed as G6PDd. Such participants had a higher post COVID-19 G6PD activity (P = 0.001; Figure 2).

**Figure 2.**
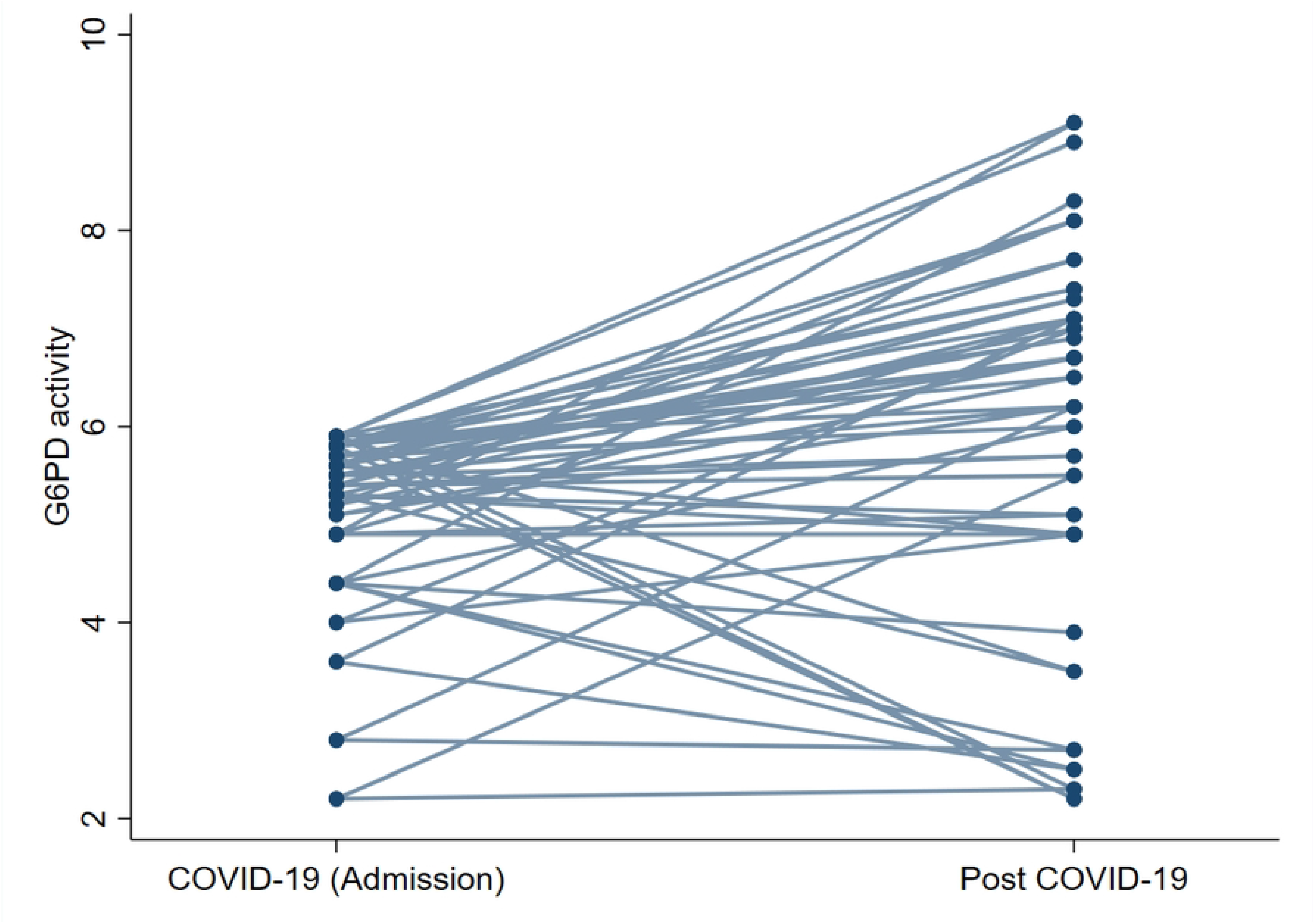
G6PD activity during and after COVID-19.

## DISCUSSION

G6PDd is the most prevalent enzyme deficiency globally and the most common form of red blood cell enzymopathy. The overall G6PDd allele frequency across malaria-endemic countries is estimated to be 8% [47]. According to Monteiro et al., in the Amazon region, prevalence ranges from 4% in the southern areas to 10% on the borders of Guyana [44]. In Manaus, the frequency of G6PDd is about 4% [45,48]. At least 186 mutations have been characterized in the G6PD gene, although not all are polymorphic and have clinical significance. In Latin America, A G6PD A-202A mutation was the variant most broadly distributed and was identified in 81.1% of the deficient individuals surveyed [44]. Santana and collaborators [49] showed the estimated frequency of the Mediterranean G6PDd (1%), and G6PDd A-(4%) variants in rural communities in Manaus. G6PD A-variant is associated with mild-to-moderate enzyme deficiency (class III) with residual enzyme activity (10–60%) and is recognized predominately in African American population [50,51]. Deficiency phenotypes are classified according to the quantification of G6PD activity, which directly impacts the severity of the hemolysis [52]. Here, G6PD activity was determined using the validated system SD Biosensor STANDARD G6PD® equipment, discriminating G6PDd from G6PDn patients and quantitatively defines the enzyme activity [53]We evidenced a much higher prevalence of G6PDd (16.3%) in patients with COVID-19 when compared to other studies carried out in Manaus.

High prevalence was also observed in a study conducted in Sardinia-Italy, showing that the frequency of G6PDd patients was higher among COVID-19 patients compared to heath population [36]. (36) Although suggesting a higher susceptibility of G6PD patients to SARS-CoV-2 infection, which has been previously demonstrated *in vitro* [54] and in an observational study [36] and expert opinion [14], our results, as well as those of Kumar et al [41]did not demonstrate an association of G6PDd with COVID-19 severity in the study population. The frequency of G6PDd patients hospitalized or demanding invasive mechanical ventilation was lower than G6PDn patients.

On admission, G6PDd patients had increased hemoglobin levels and decreased leukocytes. On the follow-up, the minimum hemoglobin values were also higher in the G6PDd group, in contrast to the maximum direct bilirubin values, which were lower than those presented by the G6PDn group. These findings diverge from previous studies, which have reported a decreased hemoglobin level, increased blood neutrophils and increased bilirubin in G6PDd patients with COVID-19 [37,55,56]. Analyzing a selected subgroup, one year later, we showed that only one-third of those participants previously classified as deficient were correctly classified, suggesting a G6PD activity modulation during the illness. Influenza virus in *in vitro* assays downregulates the expression and activity of G6PD during infection, inducing high levels of oxidative stress that favor viral replication [57]. After acute malaria, a change in G6PDd activity is also demonstrated, reinforcing that enzyme activity is influenced in acute infections [58]. Our results suggest that SARS-CoV-2 may play an influenza virus-like mechanism to increase oxidative stress and therefore, that activity is influenced by the virus.

In this study, classification of G6PD deficiency during COVID-19 was performed by quantifying enzyme activity. This diagnostic method may have skewed the results that assessed the relationship between G6PD deficiency and the severity of COVID-19. After disease resolution, participants’ enzyme activity increased, suggesting that caution should be exercised when classifying G6PD deficiency by enzyme activity during SARS-CoV-2 infection.

This study has limitations. In this study, about 20% of the participants were not confirmed by RT-PCR because laboratory confirmation was not available for all patients at that time. Manaus was facing higher levels of COVID-19 transmission which allow us to assume that SARS cases were mostly caused by SARS-Cov-2. Also, because of high mortality, we were not allowed to reach all participants for reassessing G6PD enzyme level after infection, only those who survived or allowed the procedure. Furthermore, we only reassessed those participants who were previously classified as deficient.

In conclusion, this study does not demonstrate an association of G6PDd with the severity of COVID-19. Limitations of the test for detecting enzyme levels during COVID-19 disease was demonstrated by results from both genotyping and retests carried out after the disease period showing that care must be taken when screening for G6PD deficiency in patients with acute COVID-19, and that the clinicians must evaluate the risks and benefits of different interventions, especially drugs.

## Data Availability

The data underlying the results presented in the study are available from https://www.ipccb.org/

## ACKNOWLEDGMENTS

The authors thank the CloroCovid-19 and Metcovid Team for contributing to clinical and laboratory support to patients. We also thank the participants of the clinical trials and their family members for their collaboration and support for science.

---

**Supplementary table 1.** Medications prior to inclusion in the study.

| **Variables** | **Total**<br>**N=753** | **G6PDn**<br>**N=630** | **G6PDd**<br>**N=123** | ***P*** |
| --- | --- | --- | --- | --- |
| Previous medication use, n/N (%) | 704/750 (93.9) | 587/627 (93.6) | 117/123 (95.1) | 0.530 |
| Hydrochloroquine or chloroquine in the last 30 days | 71/731 (9.7) | 62/613 (10.1) | 9/118 (7.6) | 0.400 |
| Ibuprofen | 18/702 (2.6) | 14/585 (2.4) | 4/117 (3.4) | 0.520 |
| Corticoid | 68/703 (9.7) | 60/586 (10.2) | 8/117 (6.8) | 0.260 |
| Antibiotic | 503/705 (71.3) | 426/588 (72.4) | 77/117 (65.8) | 0.150 |
| Azithromycin | 370/486 (76.1) | 315/412 (76.5) | 55/74 (74.3) | 0.690 |
| Bronchodilator | 47/705 (6.7) | 43/588 (7.3) | 4/117 (3.4) | 0.120 |
| ACE inhibitor | 222/704 (31.5) | 191/587 (32.5) | 31/117 (26.5) | 0.200 |
| Calcium Blocker | 21/704 (3.0) | 20/587 (3.4) | 1/117 (0.9) | 0.140 |
| Statin | 32/705 (4.5) | 26/588 (4.4) | 6/117 (5.1) | 0.740 |
| Immunobiological | 1/705 (0.1) | 1/588 (0.2) | 0/117 (0.0) | 0.660 |
| Antirretroviral | 9/705 (1.3) | 7/588 (1.2) | 2/117 (1.7) | 0.650 |

Abbreviations: ACE, Angiotensin-converting enzyme.

**Supplementary table 2.** Drugs used in the follow-up.

| Variables | Total<br>N=753 | G6PDn<br>N=630 | G6PDd<br>N=123 | P |
| --- | --- | --- | --- | --- |
| Medication used in studies, n/N (%) |  |  |  | 0.300 |
| Chloroquine | 59/753 (7.8) | 48/630 (7.6) | 11/123 (8.9) |  |
| Chloroquine and Antibiotic | 339/753 (45.0) | 289/630 (45.9) | 50/123 (40.7) |  |
| Antibiotic | 74/753 (9.8) | 63/630 (10.0) | 11/123 (8.9) |  |
| Control | 48/753 (6.4) | 36/630 (5.7) | 12/123 (9.8) |  |
| Drugs used in the follow-up, n/N (%) | 233/753 (30.9) | 194/630 (30.8) | 39/123 (31.7) |  |
| Antibiotic | 545/667 (81.7) | 468/565 (82.8) | 77/102 (75.5) | 0.078 |
| Azithromycin | 341/532 (64.1) | 291/459 (63.4) | 50/73 (68.5) | 0.400 |
| Ceftriaxone | 425/532 (79.9) | 363/460 (78.9) | 62/72 (86.1) | 0.160 |
| Insulin | 189/640 (29.5) | 170/545 (31.2) | 19/95 (20.0) | **0.027** |
| Corticoid | 131/668 (19.6) | 118/565 (20.9) | 13/103 (12.6) | 0.052 |
| Hydrocortisone | 69/131 (52.7) | 64/118 (54.2) | 5/13 (38.5) | 0.280 |
| Dexamethasone | 18/131 (13.7) | 16/118 (13.6) | 2/13 (15.4) | 0.860 |
| Betamethasone | 16/131 (12.2) | 15/118 (12.7) | 1/13 (7.7) | 0.600 |
| Prednisone | 11/131 (8.4) | 10/118 (8.5) | 1/13 (7.7) | 0.920 |
| Prednisolone | 7/131 (5.3) | 6/118 (5.1) | 1/13 (7.7) | 0.690 |
| Tamiflu | 374/667 (56.1) | 320/565 (56.6) | 54/102 (52.9) | 0.490 |
| Vasoactive amines | 192/668 (28.7) | 173/565 (30.6) | 19/103 (18.4) | **0.012** |
| Dopamine | 2/188 (1.1) | 2/170 (1.2) | 0/18 (0.0) | 0.640 |
| Dobutamine | 13/188 (6.9) | 12/170 (7.1) | 1/18 (5.6) | 0.810 |
| Norepinephrine | 190/191 (99.5) | 171/172 (99.4) | 19/19 (100.0) | 0.740 |
| Bronchodilators | 104/668 (15.6) | 90/566 (15.9) | 14/102 (13.7) | 0.580 |
| Aminophylline | 82/101 (81.2) | 71/87 (81.6) | 11/14 (78.6) | 0.790 |
| ACE inhibitor | 233/667 (34.9) | 196/565 (34.7) | 37/102 (36.3) | 0.760 |
| Captopril | 207/221 (93.7) | 177/187 (94.7) | 30/34 (88.2) | 0.160 |
| Anticoagulant | 348/631 (55.2) | 303/537 (56.4) | 45/94 (47.9) | 0.120 |
| Heparin | 57/343 (16.6) | 52/299 (17.4) | 5/44 (11.4) | 0.320 |
| Enoxaparin | 304/347 (87.6) | 267/302 (88.4) | 37/45 (82.2) | 0.240 |

Abbreviations: ACE, Angiotensin-converting enzyme; Control, patients who did not use chloroquine and antibiotics.

**Supplementary Figure 1.** Distribution of the mean and standard deviation of laboratory parameters throughout the follow-up. Hemoglobin (A); C-reactive protein (B); Lymphocytes (C); Total bilirubin (D); Indirect bilirubin (E); Direct bilirubin (F); Alanine aminotransferase (G); Aspartate aminotransferase (H); Lactate dehydrogenase (I); D-dimer (J).

## REFERENCES

1. Chakraborty I, Maity P. COVID-19 outbreak: Migration, effects on society, global environment and prevention. Science of the Total Environment 2020; 728.

2. Guo Y, Cao Q, Hong Z, et al. The origin, transmission and clinical therapies on coronavirus disease 2019 (COVID-19) outbreak – an update on the status. Mil Med Res 2020; :1–10.

3. Song Y, Liu P, Shi XL, et al. SARS-CoV-2 induced diarrhoea as onset symptom in patient with COVID-19. Gut 2020; 69:1143–1144.

4. Huang C, Wang Y, Li X, et al. Clinical features of patients infected with 2019 novel coronavirus in Wuhan, China. The Lancet 2020; 395:497–506.

5. Singh AK, Gupta R, Misra A. Comorbidities in COVID-19 : Outcomes in hypertensive cohort and controversies with renin angiotensin system blockers. Diabetes & Metabolic Syndrome: Clinical Research & Reviews 2020; 14:283–287.

6. Jain V, Yuan JM. Predictive symptoms and comorbidities for severe COVID-19 and intensive care unit admission: a systematic review and meta-analysis. International Journal of Public Health 2020; 65:533–546.

7. Guan W, Ni Z, Hu Y, et al. Clinical Characteristics of Coronavirus Disease 2019 in China. New England Journal of Medicine 2020; 382:1708–1720.

8. Wang D, Hu B, Hu C, et al. Clinical Characteristics of 138 Hospitalized Patients With 2019 Novel Coronavirus–Infected Pneumonia in Wuhan, China. J Am Med Assoc 2020; 323:1061–1069.

9. Tian S, Hu N, Lou J, et al. Characteristics of COVID-19 infection in Beijing. Journal of Infection 2020; 80:104–406.

10. Zhang J, Dong X, Cao Y, et al. Clinical characteristics of 140 patients infected with SARS-CoV-2 in Wuhan, China. European Journal of Allergy and Clinical Immunology 2020; 75:1730–1741.

11. Jain SK, Parsanathan R, Levine SN, Bocchini JA, Holick MF, Vanchiere JA. The potential link between inherited G6PD deficiency, oxidative stress, and vitamin D deficiency and the racial inequities in mortality associated with COVID-19. Free Radical Biology and Medicine 2020; 161:84–91.

12. Albertsen J, Ommen HB, Wandler A, Munk K. Case Report: Fatal haemolytic crisis with microvascular pulmonary obstruction mimicking a pulmonary embolism in a young African man with glucose-6-phosphate dehydrogenase deficiency. BMJ Case Reports 2014; 2014. Available at: /pmc/articles/PMC3987291/. Accessed 19 June 2022.

13. Maillart E, Leemans S, van Noten H, et al. A case report of serious haemolysis in a glucose-6-phosphate dehydrogenase-deficient COVID-19 patient receiving hydroxychloroquine. Infectious Diseases 2020; 52:659–661. Available at: https://doi.org/10.1080/23744235.2020.1774644.

14. Beauverd Y, Adam Y, Assouline B, Samii K. COVID-19 infection and treatment with hydroxychloroquine cause severe haemolysis crisis in a patient with glucose-6-phosphate dehydrogenase deficiency. European Journal of Haematology 2020; 105:357–359.

15. Franceschia L de, Costa E, Dima F, Morandi M, Olivieri O. Glucose-6-phosphate dehydrogenase deficiency associated hemolysis in COVID-19 patients treated with hydroxychloroquine/chloroquine: New case reports coming out. European Journal of Internal Medicine 2020; 80:103.

16. de Franceschi L, Costa E, Dima F, Morandi M, Olivieri O. Acute hemolysis by hydroxycloroquine was observed in G6PD-deficient patient with severe COVD-19 related lung injury. European Journal of Internal Medicine 2020; 77:136–137.

17. Sasi S, Yassin MA, Nair AP, Maslamani MSA. A case of COVID-19 in a patient with asymptomatic hemoglobin d thalassemia and glucose-6-phosphate dehydrogenase deficiency. American Journal of Case Reports 2020; 21:e925788-1-e925788-6.

18. Chaney S, Basirat A, Mcdermott R, Keenan N, Moloney E. COVID-19 & Hydroxychloroquine side-effects: Glucose 6-phosphate dehydrogenase deficiency (G6PD) and acute haemolytic anaemia. An International Journal of Medicine 2020;

19. Howes RE, Battle KE, Satyagraha AW, Hay SI. G6PD Deficiency: Global Distribution, Genetic Variants and Primaquine Therapy. Advances in Parasitology 2013; 81:133–201.

20. Howes RE, Dewi M, Piel FB, et al. Spatial distribution of G6PD deficiency variants across malaria-endemic regions. Malaria Journal 2013; 12:1–15.

21. Peters AL, Van Noorden CJF. Glucose-6-phosphate Dehydrogenase Deficiency and Malaria: Cytochemical Detection of Heterozygous G6PD Deficiency in Women. Journal of Histochemistry & Cytochemistry 2009; 57:1003–1011.

22. McMullin MF. The molecular basis of disorders of red cell enzymes. J Clin Pathol 1999; 52:241–4.

23. Christensen RD, Nussenzveig RH, Yaish HM, Henry E, Eggert LD, Agarwal AM. Causes of hemolysis in neonates with extreme hyperbilirubinemia. Journal of Perinatology 2014; 34:616–619.

24. Frank JE. Diagnosis and management of G6PD deficiency. Am Fam Physician 2005; 72:1277–82.

25. Palmer K, Dick J, French W, Floro L, Ford M. Methemoglobinemia in Patient with G6PD Deficiency and SARS-CoV-2 Infection. Emerging Infectious Diseases 2020; 26:2279–2281.

26. Mason PJ, Bautista JM, Gilsanz F. G6PD deficiency: the genotype-phenotype association. Blood Reviews 2007; 21:267–283.

27. Brito-Sousa JD, Santos TC, Avalos S, et al. Clinical Spectrum of Primaquine-induced Hemolysis in Glucose-6-Phosphate Dehydrogenase Deficiency: A 9-Year Hospitalization-based Study from the Brazilian Amazon. Clinical Infectious Diseases 2019; 69:1440–1442.

28. Brand JMA Van Den, Haagmans BL, Riel D Van, Osterhaus ADME, Kuiken T. The Pathology and Pathogenesis of Experimental Severe Acute Respiratory Syndrome and Influenza in Animal Models. Journal of Comparative Pathology 2014; 151:83–112.

29. Delgado-roche L, Mesta F. Oxidative Stress as Key Player in Severe Acute Respiratory Syndrome Coronavirus (SARS-CoV) Infection. Archives of Medical Research 2020; 51:384–387.

30. Buinitskaya Y, Gurinovich R, Wlodaver CG, Kastsiuchenka S. Centrality of G6PD in COVID-19: The Biochemical Rationale and Clinical Implications. Frontiers in Medicine 2020; 7:1–11.

31. Imai Y, Kuba K, Neely GG, et al. Identification of Oxidative Stress and Toll-like Receptor 4 Signaling as a Key Pathway of Acute Lung Injury. Cell 2008; 133:235–249.

32. Smits SL, de Lang A, van den Brand JMA, et al. Exacerbated innate host response to SARS-CoV in aged non-human primates. PLoS Pathogens 2010; 6.

33. Lin CW, Lin KH, Hsieh TH, Shiu SY, Li JY. Severe acute respiratory syndrome coronavirus 3C-like protease-induced apoptosis. FEMS Immunology and Medical Microbiology 2006; 46:375–380.

34. Kassi EN, Papavassiliou KA, Papavassiliou AG. G6PD and chloroquine: Selecting the treatment against SARS-CoV-2? Journal of Cellular and Molecular Medicine 2020; 24:4913–4914.

35. Aydemir D, Ulusu NN. Is glucose-6-phosphate dehydrogenase enzyme deficiency a factor in Coronavirus-19 (COVID-19) infections and deaths ? Pathogens and Global Health 2020; 114:109–110.

36. Littera R, Campagna M, Deidda S, et al. Human Leukocyte Antigen Complex and Other Immunogenetic and Clinical Factors Influence Susceptibility or Protection to SARS-CoV-2 Infection and Severity of the Disease Course. The Sardinian Experience. Frontiers in Immunology 2020; 11.

37. Youssef JG, Zahiruddin F, Youssef G, et al. G6PD deficiency and severity of COVID19 pneumonia and acute respiratory distress syndrome: tip of the iceberg? Annals of Hematology 2021; 100:667–673.

38. Vick DJ. Glucose-6-Phosphate Dehydrogenase Deficiency and COVID-19 Infection. Mayo Clinic Proceedings 2020; 95:1803–1804.

39. Yusuf Mohamud MF, Mukhtar MS. Epidemiological characteristics, clinical relevance, and risk factors of thromboembolic complications among patients with COVID-19 pneumonia at A teaching hospital: Retrospective observational study. Ann Med Surg (Lond) 2022; 77. Available at: https://pubmed.ncbi.nlm.nih.gov/35493413/. Accessed 19 June 2022.

40. Jalali F, Hatami F, Saravi M, et al. Characteristics and outcomes of hospitalized patients with cardiovascular complications of COVID-19. J Cardiovasc Thorac Res 2021; 13:355–363. Available at: https://pubmed.ncbi.nlm.nih.gov/35047140/. Accessed 19 June 2022.

41. Kumar N, AbdulRahman AK, AlAwadhi AI, AlQahtani M. Is glucose-6-phosphatase dehydrogenase deficiency associated with severe outcomes in hospitalized COVID-19 patients? Scientific Reports 2021; 11:19213. Available at:/pmc/articles/PMC8478975/. Accessed 19 June 2022.

42. Manaus (AM) | Cidades e Estados | IBGE. Available at: https://www.ibge.gov.br/cidades-e-estados/am/manaus.html. Accessed 20 June 2022.

43. Zobrist S, Brito M, Garbin E, et al. Evaluation of a point-of-care diagnostic to identify glucose-6-phosphate dehydrogenase deficiency in brazil. PLoS Neglected Tropical Diseases 2021; 15.

44. Monteiro WM, Val FFA, Siqueira AM, et al. G6PD deficiency in Latin America: systematic review on prevalence and variants. Memórias do Instituto Oswaldo Cruz 2014; 109:553–568.

45. Brito MAM, Peixoto HM, de Almeida ACG, et al. Validation of the rapid test Carestart™ G6PD among malaria vivax-infected subjects in the Brazilian Amazon. Rev Soc Bras Med Trop 2016; 49:446–455.

46. Hsu J, Fink D, Langer E, et al. PCR-based allelic discrimination for glucose-6-phosphate dehydrogenase (G6PD) deficiency in Ugandan umbilical cord blood. Pediatr Hematol Oncol 2014; 31:68–75. Available at: https://pubmed.ncbi.nlm.nih.gov/24308819/. Accessed 19 June 2022.

47. Howes RE, Piel FB, Patil AP, et al. G6PD Deficiency Prevalence and Estimates of Affected Populations in Malaria Endemic Countries: A Geostatistical Model-Based Map. PLoS Medicine 2012; 9.

48. Santana MS, de Lacerda MVG, Barbosa M das GV, Alecrim WD, Alecrim M das GC. Glucose-6-Phosphate Dehydrogenase Deficiency in an Endemic Area for Malaria in Manaus: A Cross-Sectional Survey in the Brazilian Amazon. PLoS ONE 2009; 4:e5259.

49. Santana MS, Monteiroa WM, Siqueiraa AM, et al. Glucose-6-phosphate dehydrogenase deficient variants are associated with reduced susceptibility to malaria in the brazilian amazon. Trans R Soc Trop Med Hyg 2013; 107:301–306.

50. Jain SK, Parsanathan R, Levine SN, Bocchini JA, Holick MF, Vanchiere JA. The potential link between inherited G6PD deficiency, oxidative stress, and vitamin D deficiency and the racial inequities in mortality associated with COVID-19. Free Radical Biology and Medicine 2020; 161:84–91.

51. Beutler E. Glucose-6-phosphate dehydrogenase deficiency. The New England Journal of Medicine 1991; 324:169–174.

52. Powers JL, Best DH, Grenache DG. Genotype-Phenotype Correlations of Glucose-6-Phosphate-Deficient Variants Throughout an Activity Distribution. J Appl Lab Med 2018; 2:841–850.

53. Zobrist S, Brito M, Garbin E, et al. Evaluation of a point-of-care diagnostic to identify glucose-6-phosphate dehydrogenase deficiency in Brazil. PLoS Negl Trop Dis 2021; 15. Available at: https://pubmed.ncbi.nlm.nih.gov/34383774/. Accessed 19 June 2022.

54. Wu YH, Tseng CP, Cheng ML, Ho HY, Shih SR, Chiu DTY. Glucose-6-phosphate dehydrogenase deficiency enhances human coronavirus 229E infection. Journal of Infectious Diseases 2008; 197:812–816.

55. Aydemir D, Daglioglu G, Candevir A, et al. COVID-19 may enhance risk of thrombosis and hemolysis in the G6PD deficient patients. Nucleosides, Nucleotides and Nucleic Acids 2021; 40:505–517.

56. Ibrahim H, Perl A, Smith D, et al. Therapeutic blockade of inflammation in severe COVID-19 infection with intravenous N-acetylcysteine. Clinical Immunology 2020; 219.

57. de Angelis M, Amatore D, Checconi P, et al. Influenza Virus Down-Modulates G6PD Expression and Activity to Induce Oxidative Stress and Promote Its Replication. Frontiers in Cellular and Infection Microbiology 2022; 11.

58. Ley B, Alam MS, Satyagraha AW, et al. Variation in Glucose-6-Phosphate Dehydrogenase activity following acute malaria. PLOS Neglected Tropical Diseases 2022; 16:e0010406. Available at: https://dx.plos.org/10.1371/journal.pntd.0010406.

